# Validity of the “HEART” Score as an Early Assessment Tool in Acute Coronary Syndrome in a Sri Lankan Population: A Single Centre Pilot Study

**DOI:** 10.1101/2025.11.05.25339585

**Authors:** C Welhenge, KM Fernando, JN Fernando, PC Deshapriya, MLD Lekamge, A Kasturiratne, AP De Silva

**Affiliations:** University Medical Unit, Colombo North Teaching Hospital, Ragama, Sri Lanka; Department of Biochemistry and Clinical Chemistry, Faculty of Medicine, University of Kelaniya, Sri Lanka; Department of Chemical Pathology, Colombo North Teaching Hospital, Ragama, Sri Lanka; Emergency department, Colombo North Teaching Hospital, Ragama, Sri Lanka; Global Health Research Unit, Department of Public Health, Faculty of Medicine, University of Kelaniya, Sri Lanka; Department of Public Health, Faculty of Medicine, University of Kelaniya, Sri Lanka; Department of Medicine, Faculty of Medicine, University of Kelaniya, Sri Lanka

## Abstract

**Objectives:** The HEART score is a clinical tool used for early risk stratification in patients presenting with chest pain in emergency settings, which facilitates prognostication. Although limited, it has been validated in several countries throughout the world. This study aimed to assess the validity of HEART score, in a Sri Lankan population, as an early assessment tool, for risk prediction of acute coronary syndrome (ACS), in patients presenting with chest pain.

**Methods:** Data was collected from 74 patients presenting to the emergency department at a tertiary care centre in Sri Lanka. HEART score was calculated for each patient retrospectively and patients were categorized into low (0-3), intermediate (4-6), and high (7-10) risk groups. The predictive accuracy of the HEART score with a diagnosis of ACS and the occurrence of major adverse cardiac events (MACE) at 6 weeks was assessed. Statistical analysis was performed using R studio.

**Results:** All patients in the low-risk group (n=8) were correctly identified as non-ACS, with no MACE. All high-risk patients (n=29) had ACS and experienced MACE. Among the intermediate risk group (n=37), 70.3% were diagnosed with ACS and 54% developed MACE at 6 weeks. Area under curve (AUC) for HEART score for a diagnosis of ACS, was 0.889 (95% CI: 0.8171-0.9609) while the AUC for occurrence of MACE was 0.9053 (95% CI: 0.8437 - 0.9669).

**Conclusions:** The HEART score is an effective early assessment tool which can be used in Sri Lankans, in prediction of the probability of ACS, and MACE within 6 weeks, in patients presenting with chest pain.

## INTRODUCTION

Chest pain accounts for majority of emergency medical admissions globally including Sri Lanka (1,2). However, only about 20% of these patients are ultimately diagnosed with acute coronary syndrome (ACS), which requires immediate treatment and inward care. Majority of the patients usually have non cardiac causes of chest pain (3). It is the responsibility of emergency care physicians to differentiate cardiac chest pain from non-cardiac causes and to exclude ACS and other life-threatening conditions.

A diagnosis of ACS relies on a combination of clinical history, dynamic electrocardiographic (ECG) changes, and cardiac biomarkers, interpreted in conjunction with clinical expertise (4). Excluding ACS based on history alone is not effective, due to, concerns with description of the chest pain, based on patient’s perception and the interpretation of the symptoms by the physician. Current guidelines recommend testing high-sensitivity cardiac troponin I (hs-cTn I) at presentation and repeating it 1–2 hours later. If the 2 hours value is normal, but the clinical suspicion is still high, a third measurement may be taken at 3 hours (5). Dynamic ECG changes with or without positive hs-cTn values confirm a diagnosis of ACS (4-6).

Delays in diagnosis of ACS, directly delay the treatment initiation, which can increase the morbidity and mortality of these patients (4, 5 – 7). These delays in diagnosis referred to as “rule out delays” contribute to overcrowding at the emergency department (ED) (8). Many patients presenting with chest pain are low risk and can be safely discharged and followed up as outpatients (9). International guidelines recommend the use of risk stratification tools in patients with chest pain due to their superiority over clinical assessment used alone and their benefits in patient outcomes (7).

The HEART score is a tool developed in 2008 for risk stratification in patients with chest pain. It is based on the five components: history, ECG characteristics, age, risk factors and troponin levels. It categorizes patients into low, intermediate or high risk, according to the short-term risk of major adverse cardiac events (MACE). These major cardiac events include acute myocardial infarction, the need for percutaneous coronary interventions (PCI) or coronary artery bypass grafting (CABG) and death within six weeks (9). Low risk patients have a reported MACE rate of 1.7% rate and can generally be safely discharged from the ED without extensive cardiac evaluation (9).

The HEART score predicts the probability of ACS with only one cardiac troponin value and thus provide the rationale for early discharge of patients with non-cardiac chest pain or patients at low risk of ACS. The use of this score has been recommended as a quick, easy to use, reliable predictor of ACS at the ED as a triage tool (1). This is particularly ideal for a low- and middle-income countries like Sri Lanka, where ED and inpatient wards frequently experience overcrowding. This pattern not only prolongs hospital stays and increases healthcare cost, but also risk compromising care for critically ill patients, due to the exhaustive burden on the health system (8, 10).

Many studies throughout the years have validated the HEART score for various populations including those in Netherland, United Kingdom, United States and South Korea (1, 11–18). However, it has not been incorporated into any formal cardiology guidelines so far. A study conducted in India has validated the HEART score for Indian population, suggesting relevance for the broader South Asian region (18). However, the score has yet to be validated for the Sri Lankan population. The purpose of this study is to assess the effectiveness of the “HEART” score at predicting the probability of ACS in patients presenting with chest pain and the effectiveness of its use at the ED as a triage tool in detecting the high-risk patients who need in ward assessment. It further attempts to assess the suitability of the HEART score as a risk assessment tool for clinical use in Sri Lankan healthcare settings.

## METHODS

### Participants

This validation study was conducted at a single tertiary care centre in Sri Lanka from July 2024 to December 2024. Data was collected from 74 adult patients presenting to the ED with chest pain. All male and female patients above 18 years complaining of chest pain who were willing to participate in the study was included. Exclusion criteria comprised pregnant and post-partum patients (up to 3 months), individuals with chest pain following accidental injuries and those with known familial heart diseases. At the time of the study, PCI facilities were not available for emergency admissions at the relevant hospital. Hence, patients diagnosed with ST-elevation myocardial infarction (STEMI) who were eligible for PCI were transferred to a specialized PCI centre and were excluded from the study. However, patients who received thrombolytic therapy were included.

### Data

An interviewer-administered electronic questionnaire was used to collect data from the patients. Additional required data was obtained from the bed head tickets (BHTs) and the hospital’s digital laboratory information system. Demographic details along with components of the clinical history were obtained from the patients and cross-verified with BHT entries. Serial ECGs were taken at the ED and the ward as required for patient management. ECG interpretations made by the attending medical officers was documented and independently verified by researchers themselves and relevant sections of the questionnaire filled. To calculate “HEART” score, the ECG characteristics of all serial ECGs taken at the ED, within 1 hour of admission was considered. hs-cTn I levels were requested from the biochemistry laboratory as a routine practice from the ED or the ward immediately upon admission. The hs-cTn I was analysed using Vitros (Ortho Clinical) 5600 analyser by Chemiluminescence immunoassay technique. This hs-cTn I value was traced from the laboratory, once available and the relevant section of the questionnaire filled.

The data collection process occurred independently of patient management and the calculated HEART scores were not disclosed to or used by the treating clinical team during decision-making. The patients were reviewed prior to discharge to assess them for the final diagnosis, which was made by the treating clinical team based on the history, examination, serial ECGs, troponin values and other relevant investigations. The required information was obtained from the BHTs and confirmed with the treating clinical team. Six-week follow-up data on the occurrence of MACE were collected from the patients and personal medical records.

### Data preparation

Data was collected to a spreadsheet and recoded for ease of statistical analysis. The HEART score was calculated from the collected independent scores using R Studio software.

### Outcome

The diagnosis of ACS was made by the treating clinical team in accordance with the 2023 European Society of Cardiology (ESC) Guidelines for the management of acute coronary syndromes and was cross checked by the research team. Based on the clinical history, ECG findings and the hs-cTn I level, ACS was further classified as unstable angina (UA), Non-ST elevated myocardial infarction (NSTEMI) and ST elevated myocardial infarction (STEMI). Occurrence of acute myocardial infarction (AMI), the need for PCI or CABG and death within six weeks of the onset of chest pain was considered as MACE. Only NSTEMI and STEMI was considered as AMI.

### Statistical analysis

Categorical data was presented using frequencies and percentages. The effectiveness of the HEART score was assessed with, sensitivity, specificity, positive predictive value (PPV), negative predictive value (NPV), and area under the receiver operating characteristic curve (AUC-ROC), as measures of predictive accuracy. Chi squared tests and Fisher’s exact tests were used to assess the statistical significance of the HEART score in prediction of, ACS and the risk of short-term MACE. All statistical analyses were performed using R Studio (Version 2025.05.0+496). Statistical significance was defined as p < 0.05 two -sided.

### Ethical approval

Ethical approval for the study was obtained from the Ethical Review Committee of the Faculty of Medicine, University of Kelaniya, Sri Lanka (FWA00013225).

## RESULTS

### Participants

Data was collected from 74 patients presenting with chest pain to the ED and followed up for 6 weeks (Figure 1).

**Figure 1.**
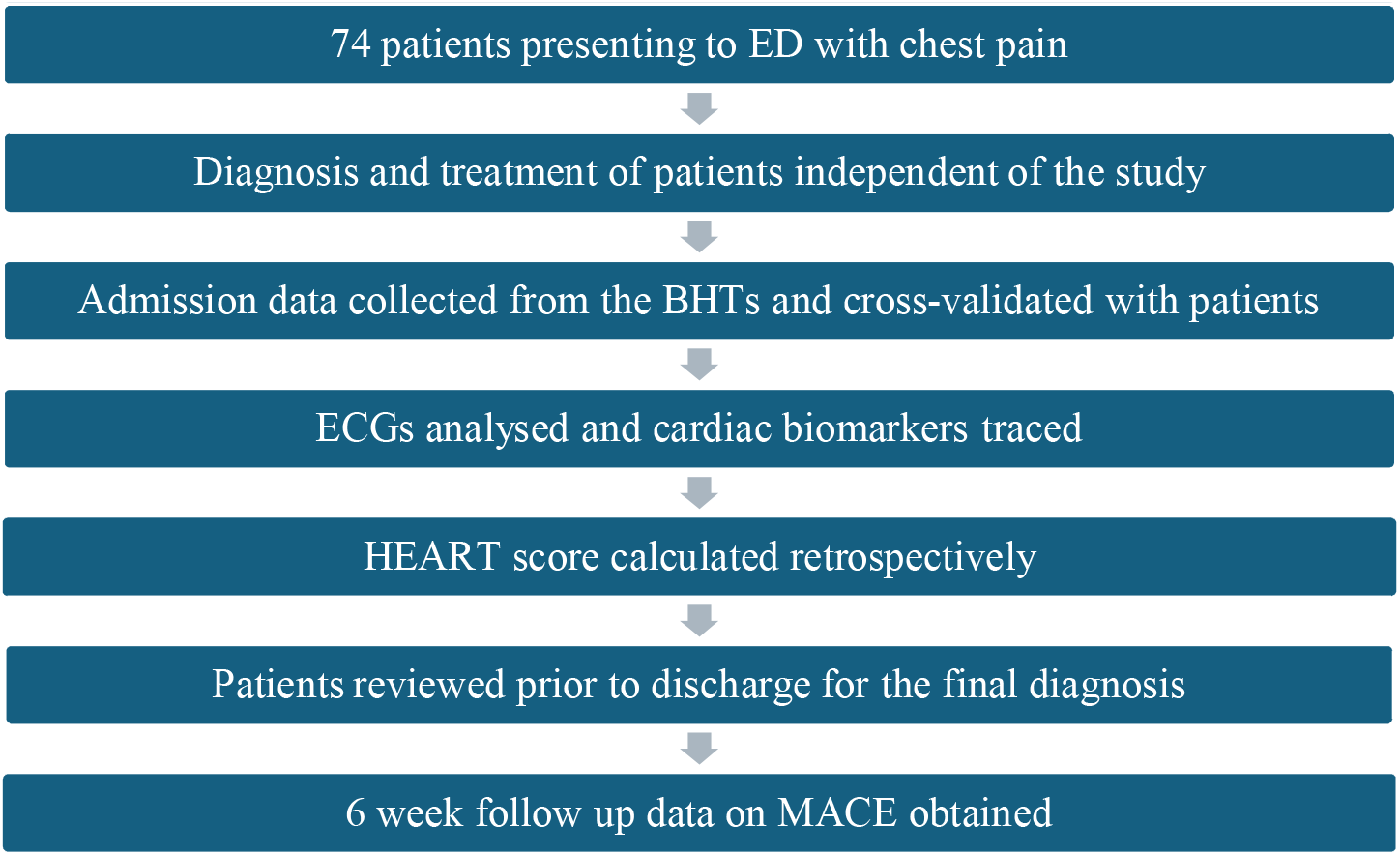
Out of the 74 patients, 42 (57%) were male and 32 (43%) were female. Patient characteristics of the study group are presented in Table 1.

**Table 1.** Patient characteristics of the study group.

| Patient Characteristics | Number |  | Percentage (%) |  |
| --- | --- | --- | --- | --- |
| Study group | 74 |  | 100 |  |
| Male | 42 |  | 57 |  |
| Female | 32 |  | 43 |  |
| Co-morbidities | ACS (55) |  | Non-ACS (19) |  |
|  | Number | Percentage (%) | Number | Percentage (%) |
| Patients with any co-morbidity | 45 | 81.8 | 12 | 63.1 |
| Diabetes | 25 | 45.5 | 4 | 21.1 |
| Smoking | 18 | 32.7 | 4 | 21.1 |
| Hypertension | 33 | 60 | 4 | 21.1 |
| Hypercholesterolemia | 16 | 29.1 | 4 | 21.1 |
| History of IHD | 25 | 45.5 | 6 | 31.6 |
| History of heart failure | 2 | 3.6 | 0 | 0 |
| Family history of CAD | 1 | 1.8 | 0 | 0 |
| History of stroke | 1 | 1.8 | 1 | 5.3 |
| CKD | 5 | 9.1 | 0 | 0 |
| Bronchial asthma | 1 | 1.8 | 2 | 10.5 |
| COPD | 1 | 1.8 | 0 | 0 |
| Hypothyroidism | 0 | 0 | 2 | 10.5 |
| Atrial fibrillation | 1 | 1.8 | 0 | 0 |
| GORD | 1 | 1.8 | 0 | 0 |
| Valvular heart disease | 0 | 0 | 1 | 5.3 |
| **Pulmonary hypertension** | 0 | 0 | 1 | 5.3 |
| **Parkinson disease** | 0 | 0 | 1 | 5.3 |
| **Thalassemia trait** | 1 | 1.8 | 0 | 0 |
| **History of tuberculosis** | 1 | 1.8 | 0 | 0 |
| **Dilated cardiomyopathy** | 2 | 3.6 | 0 | 0 |
| **None** | 10 | 18.1 | 7 | 36.8 |

Abbreviations: IHD; ischemic heart disease, CAD; coronary artery disease, CKD; chronic kidney disease, COPD; chronic obstructive pulmonary disease, GORD; Gastro-oesophageal reflux disease

A total of 60 patients (81.1%) were found to have cardiac-type chest pain, while 14 patients (18.9%) were classified as having non-cardiac chest pain. Patients diagnosed with stable angina, unstable angina, NSTEMI and STEMI were considered to have cardiac type chest pain. The characteristics of the chest pain among the study participants are presented in Table 2.

**Table 2.** Chest pain characteristics of the study group.

| **ACS (55)** |  |  | **Non-ACS (19)** |  |  |
| --- | --- | --- | --- | --- | --- |
| **Chest pain localization** | **N** | **Percentage (%)** | **Chest pain localization** | **N** | **Percentage (%)** |
| Central | 34 | 61.8 | Central | 15 | 78.9 |
| Left side | 15 | 27.3 | Left side | 4 | 21.1 |
| Right side | 5 | 9.1 |  |  |  |
| None | 1 | 1.8 |  |  |  |
| **Character of Chest pain** | **N** | **Percentage (%)** | **Character of Chest pain** | **N** | **Percentage (%)** |
| Aching type | 3 | 5.5 | Aching type | 1 | 5.3 |
| Pleuritic type | 2 | 3.6 | Pleuritic type | 2 | 10.5 |
| Tightening type | 43 | 78.2 | Tightening type | 14 | 73.7 |
| Burning type | 6 | 10.9 | Burning type | 1 | 5.3 |
| Non-specific | 1 | 1.8 | Non-specific | 1 | 5.3 |
| **Onset of Chest pain** | **N** | **Percentage (%)** | **Onset of Chest pain** | **N** | **Percentage (%)** |
| Sudden | 23 | 41.8 | Sudden | 9 | 47.4 |
| Insidious | 31 | 56.4 | Insidious | 10 | 52.6 |
| Non-specific | 1 | 1.8 |  |  |  |

| **Radiation** | **N** | **Percentage (%)** | **Radiation** | **N** | **Percentage (%)** |
| --- | --- | --- | --- | --- | --- |
| To neck/jaw | 8 | 14.5 | To neck/jaw | 5 | 26.3 |
| To left upper limb | 12 | 21.8 | To left upper limb | 7 | 36.8 |
| To right upper limb | 1 | 1.8 | To right upper limb | 1 | 5.3 |
| To bilateral upper limbs | 2 | 3.6 | To bilateral upper limbs | 1 | 5.3 |
| To back | 2 | 3.6 | To back | 2 | 10.5 |
| None | 37 | 67.3 | None | 7 | 36.8 |
| **Associations** | **N** | **Percentage (%)** | **Associations** | **N** | **Percentage (%)** |
| Autonomic symptom | 30 | 54.5 | Autonomic symptom | 7 | 36.8 |
| Exertional dyspnoea | 21 | 38.1 | Exertional dyspnoea | 6 | 31.6 |
| Orthopnoea and PND | 3 | 5.5 | Orthopnoea and PND | 0 |  |
| Palpitations | 0 |  | Palpitations | 1 | 5.3 |
| Cough | 7 | 12.7 | Cough | 5 | 26.3 |
| Wheezing | 4 | 7.3 | Wheezing | 1 | 5.3 |
| Dyspeptic symptoms | 2 | 3.6 | Dyspeptic symptoms | 2 | 10.5 |

*Abbreviations: ACS; acute coronary syndrome, PND; paroxysmal nocturnal dyspnoea*

Among the patients presenting with cardiac-type chest pain, 55 were diagnosed with ACS at the time of discharge (Table 3). ACS was excluded as the cause for chest pain based on history, ECG findings and hs-cTn I value in the remaining 19. Eleven patients were diagnosed with STEMI based on ECG findings, underwent thrombolysis and received guideline-directed medical therapy. Thirty-five patients were diagnosed with NSTEMI while nine were diagnosed with unstable angina. Both groups were treated according to established ACS guidelines.

**Table 3.**
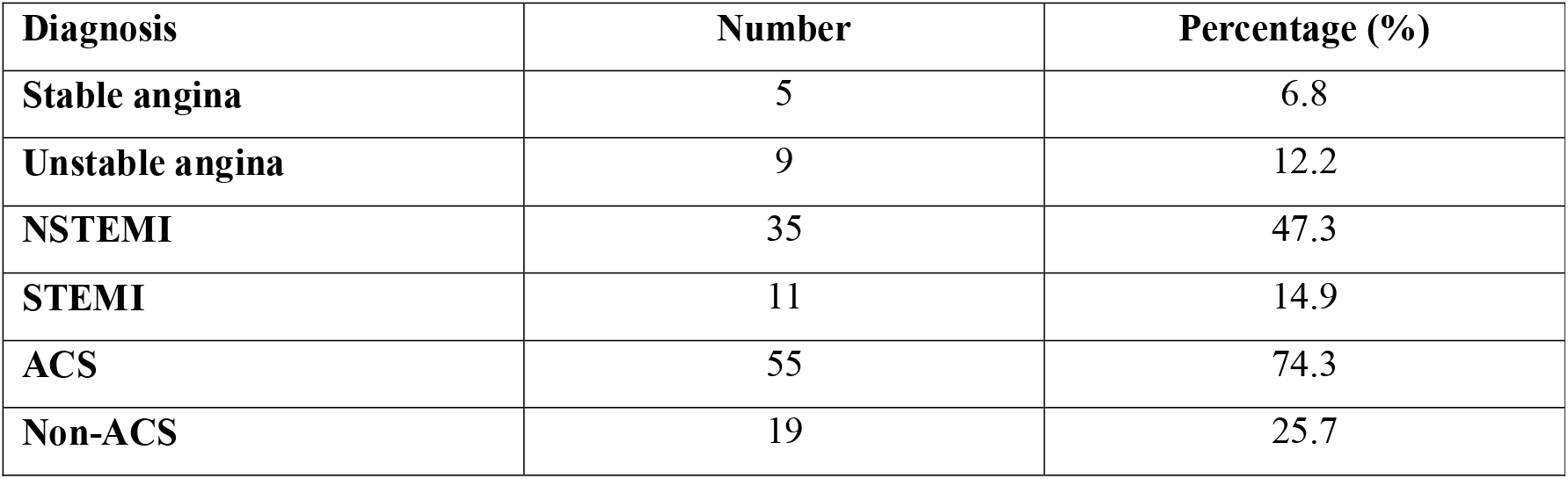
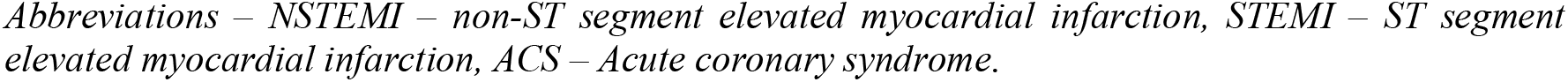
Diagnosis of patients with chest pain at discharge from the ward.

| **Diagnosis** | **Number** | **Percentage (%)** |
| --- | --- | --- |
| **Stable angina** | 5 | 6.8 |
| **Unstable angina** | 9 | 12.2 |
| **NSTEMI** | 35 | 47.3 |
| **STEMI** | 11 | 14.9 |
| **ACS** | 55 | 74.3 |
| **Non-ACS** | 19 | 25.7 |

*Abbreviations – NSTEMI – non-ST segment elevated myocardial infarction, STEMI – ST segment elevated myocardial infarction, ACS – Acute coronary syndrome.*

All patients were followed up at six weeks to assess the occurrence of MACE. Among patients diagnosed with ACS at the time of discharge, MACE was observed in 48 patients (87.3%). In contrast, only 3 patients (15.8%) in the non-ACS group experienced MACE. The types of MACE observed in each group are presented in Table 4.

**Table 4.** Occurrence of MACE at 6 weeks.

|  | ACS (55) |  | Non-ACS (19) |  |
| --- | --- | --- | --- | --- |
| **None** | 7 | 12.7% | 16 | 84.2% |
| **Myocardial infarction** | 47 | 85.5% | 3 | 15.8% |
| **PCI** | 4 | 7.3% | 1 | 5.3% |
| **CABG** | 0 |  | 0 |  |
| **Death** | 3 | 5.5% | 0 |  |
| **Total MACE** | 48 | 87.3% | 3 | 15.8% |

*Abbreviations – PCI – Percutaneous coronary interventions, CABG – Coronary artery bypass grafting, MACE – Major adverse cardiac events*

The total HEART score was calculated retrospectively for each patient and categorized into low-risk (HEART score – 0 -3), intermediate-risk (HEART score – 4 - 6) and high-risk (HEART score – 7 – 10) groups for risk stratification. Calculated HAERT scores were compared against the diagnosis at discharge. In all eight patients in the low-risk group, ACS had been excluded. All 29 patients in the high-risk group were diagnosed with ACS. Among the 37 patients in the intermediate-risk group, 26 (70.3%) were diagnosed with ACS, while ACS was excluded in 11(29.7%) (Table 5, Figure 2). Notably, all patients diagnosed with ACS (n=55) had a HEART score of intermediate or high-risk category. In intermediate and high-risk categories, HEART score was considered to be positive, while in the low-risk category it was considered to be negative. The positive predictive value (PPV) of HEART score for a diagnosis of ACS was 83.3% and the negative predictive value (NPV) was 100%.

**Table 5.** Distribution of the HEART score based on the diagnosis of ACS and occurrence of MACE.

| **HEART score risk category** | **ACS (n=55)** | **Non-ACS (n=19)** |
| --- | --- | --- |
| **Low** | 0 | 8 |
| **Intermediate** | 26 | 11 |
| **High** | 29 |  |

|  | With MACE (n=49) | Without MACE (n=25) |
| --- | --- | --- |
| **Low** | 0 | 8 |
| **Intermediate** | 20 | 17 |
| **High** | 29 | 0 |

Abbreviations – ACS – Acute coronary syndrome, MACE - Major adverse cardiac events

**Figure 2.**
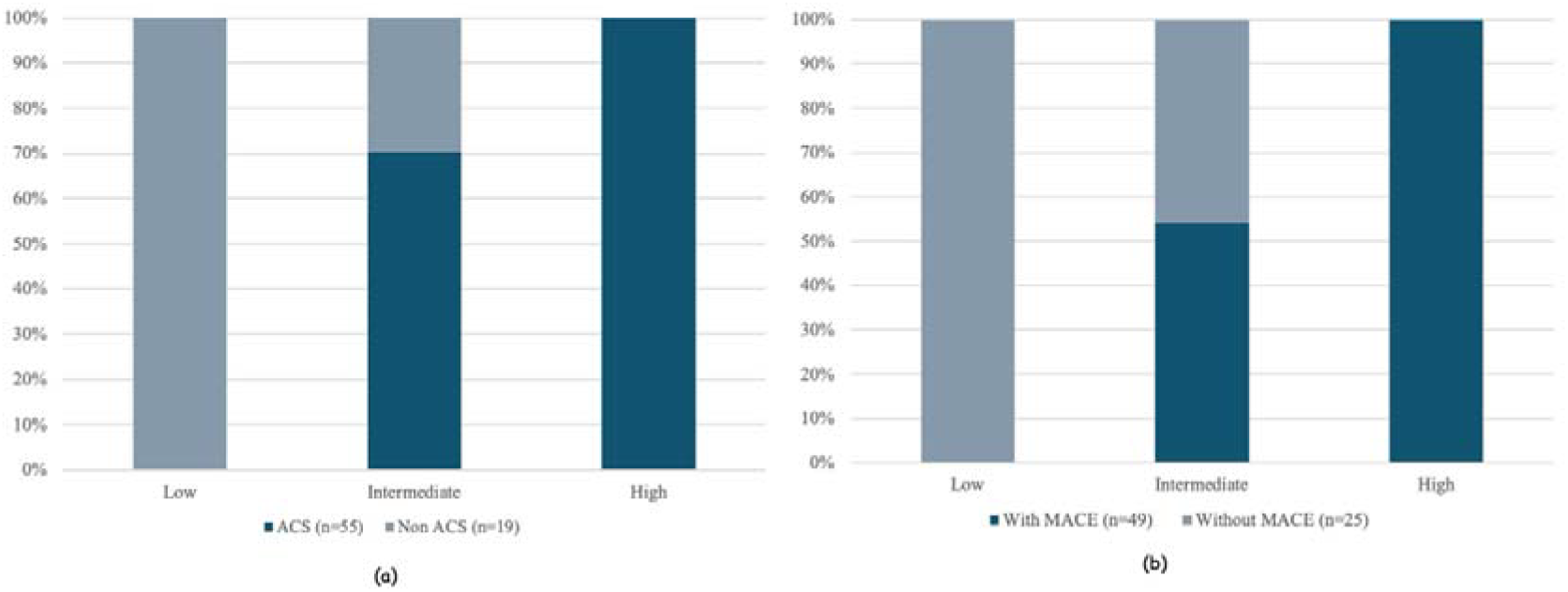
Distribution of the HEART score based on; (a) Diagnosis of ACS (b) Occurrence of MACE

An ROC curve was generated to evaluate the diagnostic performance of the HEART score for the diagnosis of ACS. The HEART score demonstrated a good diagnostic accuracy (AUC = 0.889, 95% CI: 0.8171-0.9609) (Figure 3(a)). At the optimal threshold, it yielded a high sensitivity of 72.7% and a specificity of 89.5%.

**Figure 3.**
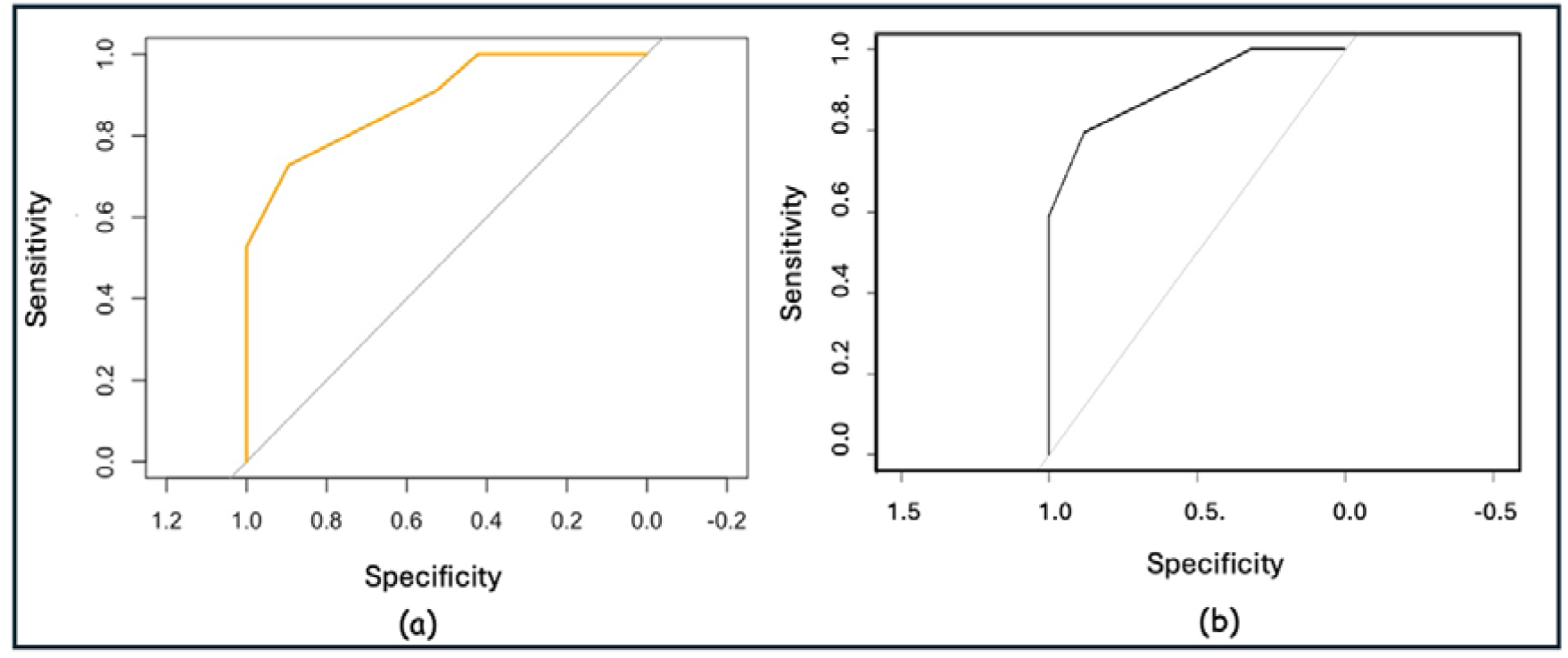
ROC curves of HEART score (a) for a diagnosis of ACS (b) for the occurrence of MACE at 6 weeks

Chi squared testing to assess the statistical significance between the ACS and non-ACS group revealed, X^2^= 33.5, df = 2, p-value – 5.33 x 10^-8^. This result was assumed to be due to the null values for low risk and high-risk categories. Fisher’s exact test revealed a p-value of 1.09 x 10^-8^, indicating a statistically significant association between HEART score risk categories and ACS diagnosis.

The HEART score was also assessed for its ability to predict MACE within six weeks. None of the patients in the low-risk group experienced MACE, while all the patients in the high-risk category experienced MACE. Among the intermediate-risk group, 20 patients (54%) experienced MACE, whereas 17 patients (45.9%) did not. Importantly, all patients who experienced MACE were classified as either intermediate- or high-risk according to their HEART scores (Table 5, Figure 2). The PPV of HEART score in predicting six week MACE was 74% while the NPV was 100%.

An additional ROC curve was generated to evaluate the HEART score’s predictive value for MACE, which demonstrated an AUC of 0.9053 (95% CI: 0.8437 - 0.9669) (Figure 3(b)), with a sensitivity of 79.6% and specificity of 88%. In keeping with previous research, this study demonstrated that there is a strong, statistically significant association between HEART score and MACE within six weeks.

Chi squared testing to assess the statistical significance between the groups with and without MACE at six weeks revealed, X^2^= 32.9, df = 2, p-value – 7.09 x 10^-8^. This result was assumed to be due to the null values for low risk and high-risk categories. Fisher’s exact test revealed a p-value of 1.965 x 10^-8^, indicating a statistically significant association between HEART score risk categories and occurrence of MACE.

## DISCUSSION

### Interpretation

Our results demonstrated that, majority of the patients presenting with chest pain, to the ED at the tertiary care hospital where the study was conducted, had cardiac type of chest pain (60/74), while only 14 had non-cardiac causes. Out of this group only patients diagnosed with ACS requires urgent inward treatment. Patients with stable angina, by contrast, could be safely discharged home after optimization of their medical treatment.

Early risk stratification of patients presenting with chest pain, using validated scores at the ED, helps the emergency physicians in rapidly ruling out ACS as well as identifying patients who require urgent medical attention. In resource limited settings like in Sri Lanka, if simple clinical tools can be utilized to predict the probability of ACS rapidly, with a single Troponin value, over-crowding at the emergency departments can be limited.

The HEART score was developed in Netherlands and had been validated in several other countries including within Asia (6,7-14) for risk stratification in patients presenting with chest pain. In our study, the HEART score demonstrated strong diagnostic performance in a Sri Lankan cohort, with an AUC of 0.889 for the diagnosis of ACS. While the PPV of HEART score for a diagnosis of ACS was 83.3%, with a sensitivity of 72.7%, the NPV of 100% with a specificity of 89.5% indicates that the score has an excellent predictive power in ruling out ACS. Further, the HEART score demonstrated excellent discriminatory power in risk prediction for six-week MACE, with an AUC of 0.9053. A sensitivity of 79.6% with a PPV of 74% and specificity of 88% and a NPV of 100% indicates that the score is best at identifying patients at low risk of MACE.

While most previous studies have primarily focussed on the prediction of 30-day or 6-week MACE in patients presenting with chest pain, we focussed on validity of the HEART score as an early assessment tool in diagnosing ACS. Although patients with unstable angina were classified under ACS, when considering the outcome as MACE, these patients were not included, hence the difference in the number with ACS and MACE.

Importantly, none of the patients, categorized as low risk by HEART score were diagnosed with ACS or experienced MACE, demonstrating the tool’s reliability in ruling out serious cardiac events in this group. Conversely, all the patients with high scores were diagnosed with ACS and developed MACE, justifying the validity of HEART score in identifying patients who require urgent care. At intermediate scores, HEART score had a better predictive accuracy for ACS rather than for MACE (ACS – 70.3%, MACE – 54%).

### Limitations

The tertiary care centre where this study was conducted was not a designated primary PCI centre at the time of data collection. Therefore, only patients who were managed within the ED and subsequently sent to the ward were included in the study. Hence the patients diagnosed with STEMI who required urgent PCI were transferred to external facilities and were not included in the study group.

Although current guidelines recommend a pharmaco-invasive strategy for patients diagnosed with STEMI, in the Sri Lankan healthcare setup, where resources are limited, not all patients who receive thrombolysis proceed to coronary angiogram routinely. In addition, CABG facilities are limited, often causing patients to wait a long time for the procedure, often beyond the study’s follow-up period. Therefore, the actual incidence of MACE might be much higher than reported in this study.

A model validation usually requires a minimum of 10-20 events (19). In our study, a total of 74 patients were included, with a MACE rate of 66% (n = 49). Despite the modest overall sample size, the high event rate exceeded the required number of outcome events to reliably assess model performance. Moreover, sample size simulations indicated that, at least 50 patients were adequate to detect clinically meaningful differences in discriminatory performance (AUC). As such, our sample size was adequate for the primary goal of external validation of an existing risk score. Nonetheless, a larger, multicentre study would enhance statistical power and improve the generalizability of the findings across the broader Sri Lankan Population.

### Usability of the model in the context of current care

Although the HEART score is already in used within emergency care set up in Sri Lanka, it has not been formally validated for the local population. This study provides evidence supporting its validity among Sri Lankans. Overcrowding is inevitable in the Sri Lankan health care setup, where resources are limited. This overcrowding can compromise the quality of patient care. By utilizing HEART score at the EDs, ACS can be ruled out rapidly in low-risk patients presenting with chest pain and hence provide the rationale for early discharge of the low-risk patients from the ED. This can be a solution for reducing the overcrowding at the ED as well as the wards.

Given the demonstrated good to excellent predictive power of the HEART score, for diagnosing ACS, it enables timely initiation of appropriate treatment for patients at risk. Considering the limited resources available, it is paramount in identifying the very high-risk group of patients with chest pain, who need to be referred for early coronary interventions, for better utilization of the limited facilities. Given the excellent discriminative power of the HAERT score to predict MACE, the HEART score can also serve as an effective communication tool between emergency physicians and cardiologists when determining urgency for referral and intervention

## Data Availability

All data produced in the present study are available upon reasonable request to the authors

## OPEN SCIENCE

### Funding

No external funding was received for the study.

### Protocol

The complete study protocol is available on request from the corresponding author.

### Registration

The study has not been registered anywhere.

### Data sharing

The data that support the findings of this study are available on request from the corresponding author.

### Code sharing

The code used for the analysis is available on request.

## PATIENT & PUBLIC INVOLVEMENT

There was no patient or public involvement in planning, design, conduct, reporting or dissemination of the study and its findings

## Abbreviations

ACS: Acute coronary syndrome
ECG: Electrocardiography
hs-cTn I: High-sensitivity cardiac troponin
I ED: Emergency department
MACE: Major adverse cardiac events
PCI: Percutaneous coronary interventions
CABG: Coronary artery bypass grafting
STEMI: ST-elevation myocardial infarction
NSTEMI: non-ST elevated myocardial
AMI: Acute myocardial infarction
PPV: Positive predictive value
NPV: negative predictive value
AUC-ROC: area under the receiver operating characteristic curve
IHD: Ischemic heart disease
CAD: Coronary artery disease
CKD: Chronic kidney disease
COPD: Chronic obstructive pulmonary disease
GORD: Gastro-oesophageal reflux disease
PND: Paroxysmal nocturnal dyspnoea

